# Clinical characteristics of 25 death cases with COVID-19: a retrospective review of medical records in a single medical center, Wuhan, China

**DOI:** 10.1101/2020.02.19.20025239

**Authors:** Xun Li, Luwen Wang, Shaonan Yan, Fan Yang, Longkui Xiang, Jiling Zhu, Bo Shen, Zuojiong Gong

## Abstract

**Background:** The pneumonia caused by the 2019 novel coronavirus (SARS-CoV-2) is a highly infectious disease, which was occurred in Wuhan, Hubei Province, China in December 2019. As of February 13, 2020, a total of 59883 cases of COVID-19 in China have been confirmed and 1368 patients have died from the disease. However, the clinical characteristics of the dyed patients were still not clearly clarified. This study aims to summarize the clinical characteristics of death cases with COVID-19 and to identify critically ill patients of COVID-19 early and reduce their mortality.

**Methods:** The clinical records, laboratory findings and radiologic assessments included chest X-ray or computed tomography were extracted from electronic medical records of 25 died patients with COVID-19 in Renmin Hospital of Wuhan University from Jan 14 to Feb 13, 2020. Two experienced clinicians reviewed and abstracted the data.

**Findings:** The mean age of the dead was 71.48 ± 12.42 years, the average course of the disease was 10.56 ± 4.42 days, all patients eventually died of respiratory failure. All of those who died had underlying diseases, the most common of which was hypertension (16/25, 64%), followed by diabetes (10/25, 40%), heart diseases (8/25, 32%), kidney diseases (5/25, 20%), cerebral infarction (4/25, 16%), chronic obstructive pulmonary disease (COPD, 2/25, 8%), malignant tumors (2/25, 8%) and acute pancreatitis (1/25, 4%). The most common organ damage outside the lungs was the heart, followed by kidney and liver. In the patients’ last examination before death, white blood cell and neutrophil counts were elevated in 17 patients (17/25, 68%) and 18 patients (18/25, 72%), lymphocyte counts were decreased in 22 patients (22/25, 88%). Most patients’ PCT, CRP and SAA levels were elevated, the percentages were 90.5% (19/21), 85% (19/20) and 100% (21/21) respectively. The levels of the last test of neutrophils (15/16, 93.8%), PCT (11/11, 100%), CRP (11/13, 84.6%), cTnI (8/9, 88.9%), D-Dimer (11/12, 91.6%) and LDH (9/9, 100%) were increased as compared to the first test, while the levels of lymphocytes were decreased (14/16, 87.5%).

**Interpretation:** The age and underlying diseases (hypertension, diabetes, etc.) were the most important risk factors for death of COVID-19 pneumonia. Bacterial infections may play an important role in promoting the death of patients. Malnutrition was common to severe patients. Multiple organ dysfunction can be observed, the most common organ damage was lung, followed by heart, kidney and liver. The rising of neutrophils, SAA, PCT, CRP, cTnI, D-Dimer and LDH levels can be used as indicators of disease progression, as well as the decline of lymphocytes counts.

## Introduction

The pneumonia caused by the 2019 novel coronavirus (SARS-CoV-2) is a highly infectious disease, which was occurred in Wuhan, Hubei Province, China in December 2019 (1). It is reported that the person-to-person transmission in hospital and family settings has been accumulating (2-4). The patients’ common clinical manifestations included fever, nonproductive cough, dyspnea, myalgia, fatigue, normal or decreased leukocyte counts, and radiographic evidence of pneumonia (5). Chen et al reported that mortality of COVID-19 was 4.3%, and severe cases (treated in the ICU) were older, more likely to have underlying comorbidities, dyspnea and anorexia (4). As of February 13, 2020, a total of 55748 cases of COVID-19 in China have been confirmed and 1380 patients have died from the disease (6). However, the clinical characteristics of the dyed patients were still not clearly clarified. In this study, we summarized the clinical characteristics of 25 death cases with COVID-19, the purpose is to identify critically ill patients of COVID-19 early and reduce their mortality.

## Methods

### Study design and patients

We performed a retrospective review of medical records from 25 death cases with COVID-19 in Renmin Hospital of Wuhan University from Jan 14 to Feb 13, 2020. All 25 dead patients with COVID-19 tested positive for severe acute respiratory syndrome coronavirus 2 (SARS-CoV-2) by use of RT-PCR on samples from there respiratory tract. Diagnosis of COVID-19 was based on the WHO’s interim guidelines(7). This study was reviewed and approved by the Medical Ethical Committee of Renmin Hospital of Wuhan University.

### Data collection

The clinical symptoms and signs, laboratory findings and radiologic assessments included chest X-ray or computed tomography were extracted from electronic medical records. Two experienced clinicians reviewed and abstracted the data. Data were entered into a computerized database and cross-checked. The criteria for the confirmed-diagnosis of SARS-CoV-2 was that at least one gene site was amplified to be positive for nucleocapsid protein (NP) gene and open reading frame (ORF) 1ab gene. In brief, the throat swab was put into a collection tube containing 150μl viral preservation solution, and the total RNA was extracted within 2h with the respiratory sample RNA separation Kit (Zhongzhi, Wuhan). The suspension was used for RT-PCR assay of SARS-CoV-2 RNA. Two target genes, including NP and ORF1ab, were simultaneously amplified and tested during the real-time RT-PCR assay. Target 1 (NP): forward primer GGGGAACTTCTCCTGCTAGAAT; reverse primer CAGACATTTTGCTCTC AAGCTG; and the probe 5’-FAM-TTGCTGCTGCTTGACAGATT-TAMRA-3’. Target 2 (ORF1ab): forward primer CCCTGTGGGTTTTACACTTAA; reverse primer ACGATTGTGC ATCAGCTGA; and the probe 5’-VIC-CCGTCTGCGGTATGTGGAAAGGTTATGG-BHQ1 −3’. The real-time RT-PCR assay was performed using a 2019-nCoV nucleic acid detection kit according to the manufacturer’s protocol (Shanghai bio-germ Medical Technology Co Ltd). Specific primers and probes for SARS-CoV-2 RNA detection were based on the recommendation by the National Institute for Viral Disease Control and Prevention (China) (http://ivdc.chinacdc.cn/kyjz/202001/t20200121_211337.html).

### Statistical analysis

Statistical analysis was done with SPSS, version 20.0. Continuous variables were directly expressed as mean ± SD. Categorical variables were expressed as number (%).

## Results

### General clinical characteristics

Of the 25 deaths, 10 were male and 15 were female. The mean age of the dead was 71.48 ± 12.42 years, range from 55 to100 years. The average course of the disease was 10.56 ± 4.42 days, range from 4 to 20 days. All patients eventually died of respiratory failure and respirator was used in 23 patients (23/25, 92%). All (25/25, 100%) of those who died had underlying diseases, the most common of which was hypertension (16/25, 64%), followed by diabetes (10/25, 40%), heart diseases (8/25, 32%), kidney diseases (5/25, 20%), cerebral infarction (4/25, 16%), chronic obstructive pulmonary disease (COPD, 2/25, 8%), malignant tumors (2/25, 8%) and acute pancreatitis (1/25, 4%). (Table.1)

**Table.1.**
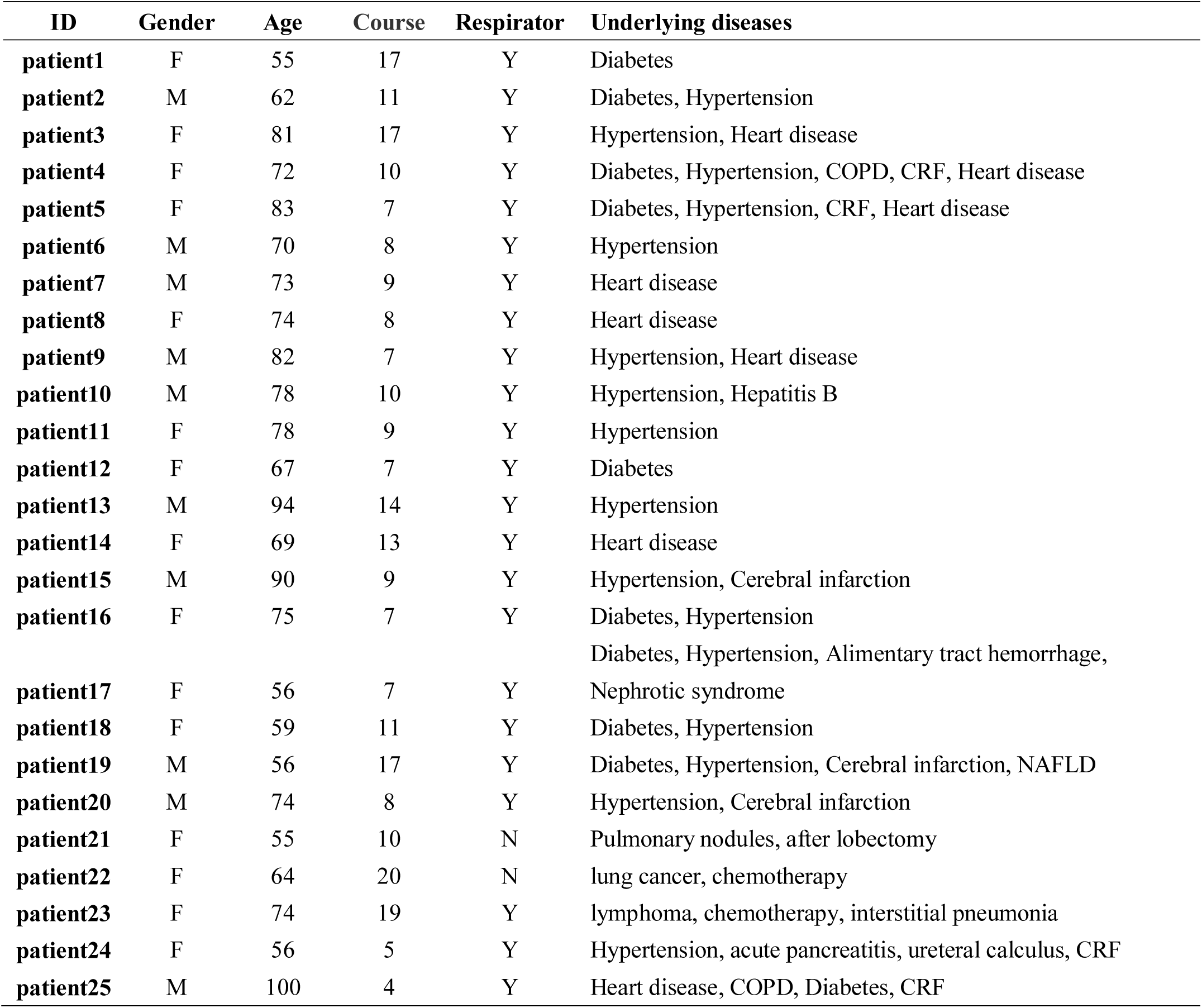
General clinical characteristics of 25 death cases.

| **ID** | **Gender** | **Age** | **Course** | **Respirator** | **Underlying diseases** |
| --- | --- | --- | --- | --- | --- |
| **patient1** | F | 55 | 17 | Y | Diabetes |
| **patient2** | M | 62 | 11 | Y | Diabetes, Hypertension |
| **patient3** | F | 81 | 17 | Y | Hypertension, Heart disease |
| **patient4** | F | 72 | 10 | Y | Diabetes, Hypertension, COPD, CRF, Heart disease |
| **patient5** | F | 83 | 7 | Y | Diabetes, Hypertension, CRF, Heart disease |
| **patient6** | M | 70 | 8 | Y | Hypertension |
| **patient7** | M | 73 | 9 | Y | Heart disease |
| **patient8** | F | 74 | 8 | Y | Heart disease |
| **patient9** | M | 82 | 7 | Y | Hypertension, Heart disease |
| **patient10** | M | 78 | 10 | Y | Hypertension, Hepatitis B |
| **patient11** | F | 78 | 9 | Y | Hypertension |
| **patient12** | F | 67 | 7 | Y | Diabetes |
| **patient13** | M | 94 | 14 | Y | Hypertension |
| **patient14** | F | 69 | 13 | Y | Heart disease |
| **patient15** | M | 90 | 9 | Y | Hypertension, Cerebral infarction |
| **patient16** | F | 75 | 7 | Y | Diabetes, Hypertension |
| **patient17** | F | 56 | 7 | Y | Diabetes, Hypertension, Alimentary tract hemorrhage, Nephrotic syndrome |
| **patient18** | F | 59 | 11 | Y | Diabetes, Hypertension |
| **patient19** | M | 56 | 17 | Y | Diabetes, Hypertension, Cerebral infarction, NAFLD |
| **patient20** | M | 74 | 8 | Y | Hypertension, Cerebral infarction |
| **patient21** | F | 55 | 10 | N | Pulmonary nodules, after lobectomy |
| **patient22** | F | 64 | 20 | N | lung cancer, chemotherapy |
| **patient23** | F | 74 | 19 | Y | lymphoma, chemotherapy, interstitial pneumonia |
| **patient24** | F | 56 | 5 | Y | Hypertension, acute pancreatitis, ureteral calculus, CRF |
| **patient25** | M | 100 | 4 | Y | Heart disease, COPD, Diabetes, CRF |

### Organ dysfunction in COVID-19

In addition to affecting respiratory function, the most common organ damage outside the lungs was the heart, (18 patients’ serum hypersensitive troponin I (cTnI) or/and Amino-terminal pro-brain natriuretic peptide (Pro-BNP) levels were increased (18/19, 94.7%)), followed by kidney (12 patients’ serum blood urea nitrogen (BUN) or/and creatinine (Cr) levels were increased (12/25, 48%)) and liver (5 patients’ serum Alanine transaminase (ALT) and aspartate aminotransferase (AST) levels were increased (5/25, 20%)). Besides, all the patients’ albumin levels were decreased. (Table.2)

**Table.2.** Changes of organ function of 25 death cases.

| **ID** | **ALT**<br>**(U/L)** | **AST**<br>**(U/L)** | **ALB**<br>**(g/L)** | **BUN**<br>**(mmol/L)** | **Cr**<br>**(μmol/L)** | **cTnI**<br>**(ng/mL)** | **Pro-BNP**<br>**(pg/mL)** |
| --- | --- | --- | --- | --- | --- | --- | --- |
| patient1 | 16 | 30 | 26.18 | 7.81 | 43 | 0.205 | 8304 |
| patient2 | 49 | 67 | 27.91 | 33.43 | 428 | 16.342 | 7961 |
| patient3 | 22 | 34 | 26.8 | 7.22 | 47 | 0.789 | 8023 |
| patient4 | 7 | 18 | 29.74 | 18.07 | 497 | 0.057 | 6914 |
| patient5 | 27 | 56 | 28.6 | 20.75 | 209 | 4.129 | 5725 |
| patient6 | 70 | 107 | 28.51 | 14.36 | 113 | 0.316 | 1182 |
| patient7 | 31 | 53 | 34.58 | N/A | N/A | N/A | 429 |
| patient8 | 14 | 34 | 35.98 | 6.07 | 58 | N/A | 90 |
| patient9 | 33 | 38 | 31.7 | 9.29 | 63 | N/A | 1658 |
| patient10 | 51 | 81 | 35.19 | 12.67 | 110 | 5.42 | 940 |
| patient11 | 21 | 38 | 37.86 | N/A | 242 | N/A | N/A |
| patient12 | 27 | 36 | 36.4 | 5.57 | 51 | 5.804 | 2450 |
| patient13 | 44 | 62 | 36.5 | 8.66 | 73 | 0.225 | 2394 |
| patient14 | 24 | 27 | 29.9 | 5.14 | 51 | N/A | N/A |
| patient15 | 17 | 43 | 34.4 | 4.89 | 60 | N/A | N/A |
| patient16 | 21 | 37 | 36.1 | 9.73 | 49 | 0.034 | 2866 |
| patient17 | 16 | 39 | 23.8 | 23.92 | 69 | 0.13 | 12733 |
| patient18 | 68 | 32 | 28.7 | 6.67 | 43 | N/A | N/A |
| patient19 | 515 | 1082 | 32.81 | 16.4 | 175 | 92.43 | 18019 |
| patient20 | 48 | 59 | 33.4 | 10.21 | 71 | 0.017 | 822 |
| patient21 | 26 | 32 | 36.1 | 2.9 | 51 | N/A | N/A |
| patient22 | 20 | 20 | 33.8 | 7.59 | 26 | 0.006 | 230 |
| patient23 | 13 | 24 | 32.5 | 1.8 | 29 | N/A | N/A |
| patient24 | 17 | 23 | 28.5 | 18.53 | 580 | 1.364 | N/A |
| patient25 | 13 | 29 | 37.16 | 9.42 | 119 | N/A | N/A |
| Normal |  |  |  |  |  |  | <75y:0-125 |
| range | 9-50 | 15-40 | 40-55 | 3.6-9.5 | 57-111 | 0-0.04 | >75y:0-450 |

N/A: Not available

### Inflammatory response in COVID-19

The routine blood test, procalcitonin (PCT), C-Reactive Protein (CRP) and Serum amyloid A (SAA) were used to reflect changes of inflammatory response in COVID-19. In the patients’ last examination before death, white blood cell and neutrophil counts were elevated in 17 patients (17/25, 68%) and 18 patients (18/25, 72%), lymphocyte counts were decreased in 22 patients (22/25, 88%). Most patients had mild anemia, RBC and Hb levels were decreased in 20 (20/25, 80%) and 17 (17/25, 68%) patients respectively. Due to alimentary tract hemorrhage, the patient 17 had severe anemia. Most patients’ PCT, CRP and SAA levels were elevated, the percentages were 90.5% (19/21), 85% (19/20) and 100% (21/21) respectively. (Table.3)

**Table.3.** Changes of inflammatory markers of 25 death cases.

| **ID** | **WBC**<br>**(cells/L)** | **NEU**<br>**(cells/L)** | **LYM**<br>**(cells/L)** | **RBC**<br>**(cells/L)** | **Hb**<br>**(g/L)** | **PLT**<br>**(cells/L)** | **PCT**<br>**(ng/mL)** | **CRP**<br>**(mg/L)** | **SAA**<br>**(mg/L)** |
| --- | --- | --- | --- | --- | --- | --- | --- | --- | --- |
| patient1 | 10.84 | 10.41 | 0.31 | 3.64 | 112 | 194 | 1.43 | 86 | >300 |
| patient2 | 17.91 | 17.01 | 0.23 | 3.92 | 137 | 39 | 75 | 96.2 | >300 |
| patient3 | 14.04 | 13.72 | 0.13 | 3.79 | 122 | 125 | 1.33 | 174 | >300 |
| patient4 | 10.08 | 9 | 0.52 | 2.44 | 68 | 167 | 3.55 | 178 | >300 |
| patient5 | 2,98 | 2.63 | 0.26 | 2.77 | 91 | 64 | N/A | 82.8 | >300 |
| patient6 | 16.74 | 15.07 | 0.28 | 3.83 | 128 | 226 | 2.09 | 203 | >300 |
| patient7 | 10.27 | 8.9 | 0.69 | 4.56 | 140 | 233 | N/A | 124.3 | >300 |
| patient8 | 8.68 | 7.48 | 0.72 | 3.97 | 120 | 224 | 0.178 | 141 | >300 |
| patient9 | 11.76 | 10.98 | 0.41 | 5.45 | 174 | 262 | 0.228 | N/A | N/A |
| patient10 | 13.93 | 12.41 | 1.12 | 4.05 | 134 | 175 | 0.376 | 180 | >300 |
| patient11 | 9.58 | 8.13 | 0.81 | 2.65 | 77 | 338 | N/A | N/A | N/A |
| patient12 | 12.61 | 10.76 | 1.09 | 4.58 | 139 | 139 | 0.126 | 51.4 | >300 |
| patient13 | 5.63 | 4.92 | 0.53 | 3.36 | 106 | 150 | 0.139 | 120 | >300 |
| patient14 | 11.01 | 10.31 | 0.55 | 3.69 | 115 | 24 | N/A | 110 | >300 |
| patient15 | 5.65 | 4,56 | 0.59 | 4.02 | 130 | 128 | 0.04 | 33.8 | 202.41 |
| patient16 | 13.35 | 12.62 | 0.52 | 4.69 | 150 | 239 | 0.12 | 39 | >300 |
| patient17 | 18.77 | 17.31 | 1.1 | 1.45 | 45 | 133 | 1.98 | 74 | >300 |
| patient18 | 6.33 | 5.39 | 0.48 | 4.2 | 123 | 139 | 0.08 | 7.6 | 73.75 |
| patient19 | 9.53 | 9.25 | 0.16 | 2.6 | 84 | 146 | 1.12 | N/A | >300 |
| patient20 | 20.17 | 17.92 | 1.43 | 4.49 | 139 | 201 | 0.3 | N/A | N/A |
| patient21 | 3.98 | 2.99 | 0.68 | 3.82 | 121 | 121 | 0.16 | 148.1 | >300 |
| patient22 | 12.7 | 12.19 | 0.14 | 3.98 | 128 | 120 | 0.11 | 68 | >300 |
| patient23 | 2.62 | 2 | 0.17 | 2.67 | 87 | 92 | 0.36 | 80.7 | >300 |
| patient24 | 17.69 | 16.36 | 0.53 | 3.78 | 101 | 200 | 1.83 | N/A | N/A |
| patient25 | 2.25 | 1.81 | 0.27 | 3.69 | 115 | 152 | 3.49 | 41.6 | 144.16 |
|  | 3.5-9.5 | 1.8-6.3 | 1.1-3.2 | 4.3-5.8 |  | 125-350 |  |  |  |
| **Normal range** | ($\times 10^9$) | ($\times 10^9$) | ($\times 10^9$) | ($\times 10^{12}$) | 130-175 | ($\times 10^9$) | 0-0.1 | 0-10 | 0-10 |

WBC: white blood cells, NEU: neutrophils, LYM: lymphocytes, RBC: red blood cells, Hb: hemoglobin, PLT: blood platelet. The detection upper limit of SAA is 300 mg/L. N/A: Not available.

**Table.4.** Specific biomarker that indicating poor prognosis.

| ID | NEU<br>(cells/L) |  | LYM<br>(cells/L) |  | PCT<br>(ng/mL) |  | CRP<br>(mg/mL) |  | SAA<br>(mg/mL) |  | cTnI<br>(ng/mL) |  | D-Dime<br>(mg/L) |  | LDH<br>(U/L) |  |
| --- | --- | --- | --- | --- | --- | --- | --- | --- | --- | --- | --- | --- | --- | --- | --- | --- |
|  | First | Last | First | Last | First | Last | First | Last | First | Last | First | Last | First | Last | First | Last |
| patient1 | 4.39 | 10.41 | 0.35 | 0.31 | 0.02 | 1.44 | 14.1 | 86 | >300 | >300 | 0.015 | 0.205 | 0.49 | 56.47 | 401 | 631 |
| patient2 | 7.27 | 17.01 | 0.32 | 0.23 | 1.42 | 75 | 83.5 | 96.2 | >300 | N/A | 0.091 | 16.34 | 4.39 | 115.3 | 758 | N/A |
| patient3 | 1.29 | 13.72 | 0.49 | 0.13 | 0.13 | 1.33 | 52.9 | 174 | 5 | >300 | 0.071 | 0.293 | 35.56 | 49.52 | 331 | 604 |
| patient4 | 8.9 | 9 | 0.9 | 0.52 | 0.89 | 3.55 | 211 | 178 | >300 | >300 | 0.075 | 0.057 | 2.82 | 1.53 | 248 | 323 |
| patient5 | 4.07 | 2.63 | 0.2 | 0.26 | N/A | N/A | 76.1 | 0.5 | >300 | >300 | 0.538 | 4.129 | N/A | N/A | N/A | N/A |
| patient6 | 7.95 | 15.07 | 0.51 | 0.28 | N/A | 2.09 | 201 | 203 | >300 | >300 | N/A | 0.316 | N/A | 20.33 | N/A | N/A |
| patient13 | 3.7 | 4.92 | 1.4 | 0.53 | 0.07 | 0.139 | 58 | 120 | 253 | >300 | 0.177 | 0.225 | 0.35 | 13.9 | 294 | 535 |
| patient14 | 4.74 | 10.31 | 0.84 | 0.55 | 0.02 | N/A | 19.9 | 110 | >300 | >300 | N/A | N/A | 1.31 | 49.8 | 397 | 510 |
| patient16 | 11.52 | 12.62 | 0.92 | 0.52 | 0.12 | N/A | 39 | N/A | >300 | N/A | 0.34 | 0.34 | 1.04 | 3.59 | 455 | N/A |
| patient17 | 14.49 | 17.31 | 1.2 | 1.1 | 0.24 | 1.98 | 56.8 | 74 | >300 | >300 | 0.006 | 0.13 | 20.57 | 1.16 | 256 | N/A |
| patient18 | 2.5 | 5.39 | 0.54 | 0.48 | 0.08 | 0.11 | 7.6 | 18.4 | 73.75 | >300 | N/A | N/A | 0.43 | 5.95 | 237 | 478 |
| patient19 | 2.89 | 9.25 | 0.7 | 0.16 | 0.11 | 1.12 | 4.9 | N/A | 11.11 | >300 | 0.06 | 92.43 | 0.14 | 155 | 252 | 669 |
| patient21 | 2.02 | 2.99 | 0.76 | 0.68 | 0.07 | 0.16 | 31.8 | 148.1 | >300 | >300 | N/A | N/A | N/A | 1.33 | N/A | 278 |
| patient22 | 9.08 | 12.19 | 0.06 | 0.14 | 0.09 | 0.11 | 52 | 32.7 | >300 | 187.8 | 0.006 | N/A | 3 | 2.33 | 321 | 322 |
| patient23 | 8.88 | 2 | 0.26 | 0.17 | 0.13 | 0.36 | 19.2 | 80.7 | 176.3 | >300 | N/A | N/A | 0.41 | 4.12 | 347 | 405 |
| patient24 | 8.08 | 16.36 | 1.07 | 0.53 | N/A | 1.83 | N/A | N/A | N/A | N/A | N/A | N/A | N/A | 13.09 | N/A | N/A |
|  | 1.8-6.3 |  | 1.1-3.2 |  |  |  |  |  |  |  |  |  |  |  |  |  |
| Normal range | ×10<sup>9</sup> |  | ×10<sup>9</sup> |  | 0-0.1 |  | 0-10 |  | 0-10 |  | 0-0.04 |  | 0-0.55 |  | 120-250 |  |

NEU: neutrophils, LYM: lymphocytes, N/A: Not available.

### Specific biomarker that indicating poor prognosis

In order to screen for biomarker indicating poor prognosis, we observed the changes of biochemical indicators in all patients (if repeated measurements were present). The results showed that the levels of the last test of neutrophils (15/16, 93.8%), PCT (11/11, 100%), CRP (11/13, 84.6%), cTnI (8/9, 88.9%), D-Dimer (11/12, 91.6%) and lactate dehydrogenase (LDH) (9/9, 100%) were increased as compared to the first test, while the levels of lymphocytes were decreased (14/16, 87.5%). The SAA maintained a high level. Chest CT scan showed that the patients’ pulmonary lesions were worse in the late stage than in the early stage (patient 3, patient 13 and patient 14). (Fig.1)

**Fig.1.**
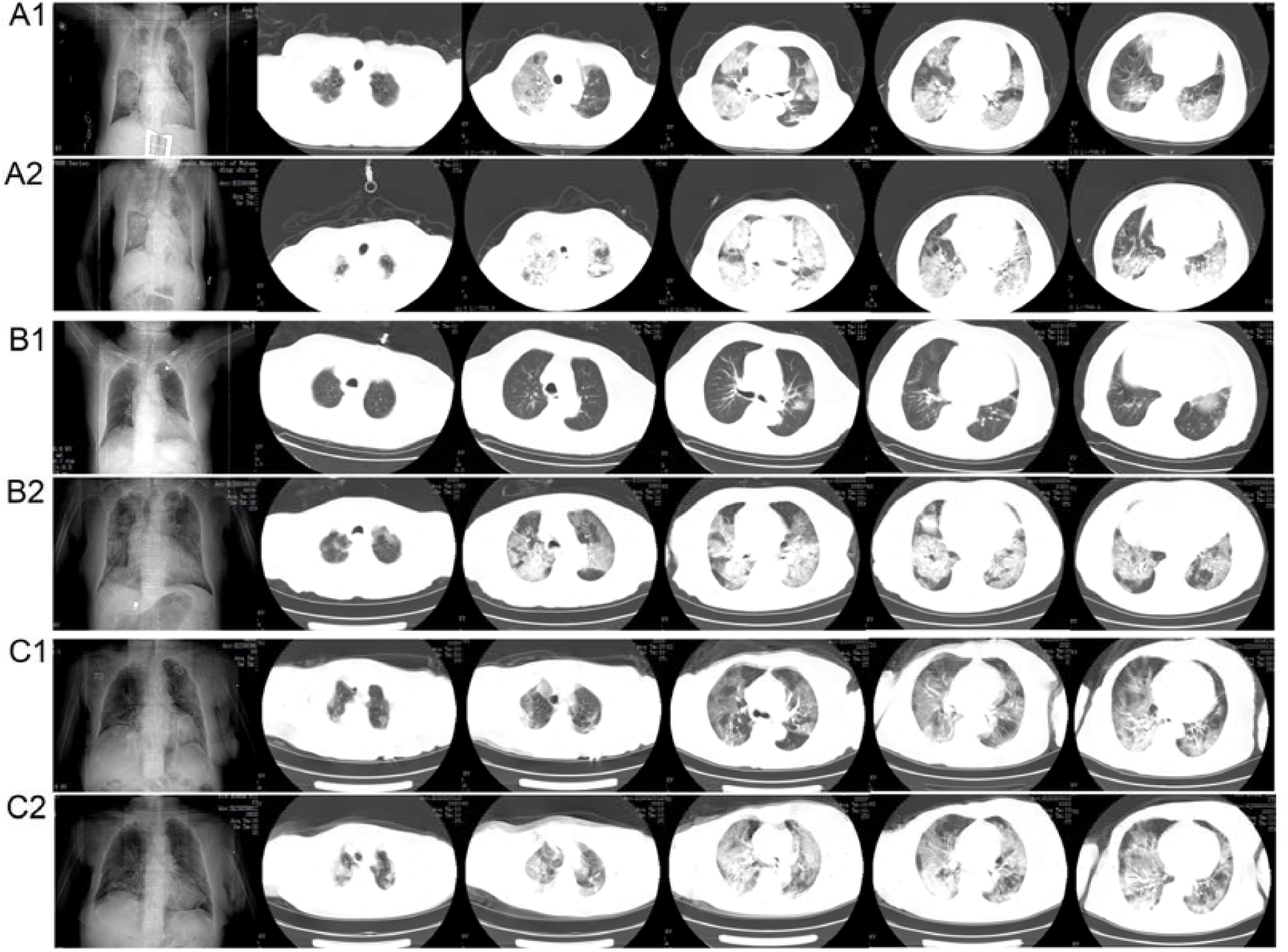
Chest CT scans of patient 3, patient 13 and patient 14. A1: the early stage Chest CT scan of Patient 3; A2: the late stage Chest CT scan of Patient 3; B1: the early stage Chest CT scan of Patient 13; B2: the late stage Chest CT scan of Patient 13; C1: the early stage Chest CT scan of Patient 14; C2: the late stage Chest CT scan of Patient 14.

## Discussion

In this study, we reported 25 death cases of with COVID-19. The clinical characters of these patients indicated that the age and underlying diseases were the most important risk factors for death. As concerning the underlying diseases, the most common one was hypertension, followed by diabetes, Heart disease, kidney disease, cerebral infarction, COPD, malignant tumors and acute pancreatitis.

The SARS-CoV-2 has been identified as one of a class of single-stranded enveloped 39 RNA viruses, belonging to the beta-coronaviruses genus of the *Coronaviridae* family (1). The analysis showed that both SARS-CoV-2 and the SARS-CoV shared a common ancestor that resembles the bat coronavirus HKU9-1 (8). And the severity of some cases with SARS-CoV-2 was similar to that of SARS-CoV (9). In the presents study, all the patients were died of respiratory failure, which indicated that the lung is the most common target organ of SARS-CoV-2.

Multiple organ dysfunction could also be observed, the most common organ damage outside the lungs was the heart, followed by kidney and liver. The results demonstrated that the death of the patient may be primarily related to impaired cardiopulmonary function. All the patients’ albumin levels and 80% and 68% of patients’ RBC and Hb levels were decreased, which indicates that malnutrition is common to severe patients.

COVID-19 is a viral disease characterized by normal or low white blood cell count and decreased lymphocyte count (10). In this study, increased white blood cell and neutrophils count were observed in 68% and 72% of patients. In addition, PCT levels were elevated in 90.5% of patients. PCT is sensitive indicator of bacterial infection (11). The results indicated that bacterial infections may play an important role in promoting the death of patients.

CRP is a useful marker and gauge of inflammation, it plays an important role in host defense against invading pathogens as well as in inflammation (12). SAA is a plasma protein that transports lipids during inflammation (13). In the present study, CRP and SAA were elevated before death in 85% and 100% of patients, suggesting that there is a severe inflammatory cascade in patients with COVID-19.

In order to screen out biochemical indicators that are meaningful for the diagnosis of disease progression, we consulted the laboratory test results of all the dead patients, among which 16 patients had repeated measurements. The SAA maintained a high level in all the patients, this result indicated that elevated SAA levels are closely related to the poor prognosis of patients. The levels of the first test of neutrophils (93.8%), PCT (100%), CRP (84.6%), cTnI (88.9%), D-Dimer (91.6%) and LDH (100%) were increased as compared to the last test, while the levels of lymphocytes were decreased (87.5%), suggesting that the rising of neutrophils, PCT, CRP, cTnI, D-Dimer and LDH levels can be used as indicators of disease progression, as well as the decline of lymphocytes counts.

This was a small sample size retrospective study, which was limited by the small numbers of patients and by using a retrospective method. In particular, some important laboratory results were incomplete.

In conclusion, the age and underlying diseases (hypertension, diabetes, etc.) is the most important risk factors for death of COVID-19. Bacterial infections may play an important role in promoting the death of patients. Malnutrition is common to severe patients. Multiple organ dysfunction can be observed, the most common organ damage outside the lungs is the heart, followed by kidney and liver. The rising of neutrophils, SAA, PCT, CRP, cTnI, D-Dimer and LDH levels can be used as indicators of disease progression, as well as the decline of lymphocytes counts.

## Data Availability

The data used to support the findings of this study are available from the corresponding author upon request.

## Contributors

ZG and BS made substantial contributions to the study concept and design. XL was in charge of the manuscript draft. LW took responsibility for obtaining ethical approval and collecting samples. FY and JZ made substantial contributions to data acquisition, analysis and interpretation. SY and LX reviewed the data. ZG made substantial revisions to the manuscript.

